# INSIGHT: A Tool for Fit-for-Purpose Evaluation and Quality Assessment of Observational Data Sources for Real World Evidence on Medicine and Vaccine Safety

**DOI:** 10.1101/2023.10.30.23297753

**Authors:** Vjola Hoxhaj, Constanza L. Andaur Navarro, Judit Riera-Arnau, Roel JHJ Elbers, Ema Alsina, Caitlin Dodd, Miriam CJM Sturkenboom

## Abstract

**Objective:** To describe the development of INSIGHT, a real-world data quality tool to assess completeness, consistency, and fitness-for-purpose of observational health data sources.

**Material and Methods:** We designed a three-level pipeline with data quality assessments (DQAs) to be performed in ConcePTION Common Data Model (CDM) instances. The pipeline has been coded using R.

**Results:** INSIGHT is an open-source tool that identifies potential data quality issues in CDM-standardized instances through the systematic execution and summary of over 588 configurable DQAs. Level 1 focuses on compliance with the ConcePTION CDM specifications. Level 2 evaluates the temporal plausibility of events and uniqueness of records. Level 3 provides an overview of distributions, outliers, and trends over time. The DQAs are run locally and assessed centrally by a data quality revisor together with the data access provider’s representatives.

**Discussion:** NSIGHT aligns with recent conceptual frameworks that identify five dimensions of data quality: reliability, extensiveness, coherence, timeliness, and relevance. Data quality is the sum of several internal and external features of the data and while DQAs provide reassurance about fitness-for-purpose for secondary-use data sources, improvements in data collection and generation stages are essential to reduce bias, misclassification, and measurement errors, thereby enhance overall data quality for Real World Evidence.

**Conclusion:** INSIGHT aims to support clinical and regulatory decision-making for medicines and vaccines by evaluating the quality of observational health data sources to support fit for purpose assessment. Assessing and improving data quality will enhance the reliability and quality of the generated evidence.

## INTRODUCTION

Regulatory agencies and healthcare professionals acknowledge and underline the significance of Real-World Data (RWD) in informing clinical decisions and shaping public health policies, especially for populations underrepresented in clinical trials, such as pregnant women.^1,2^ Pharmacoepidemiologists have used RWD since the 90s, and worked in distributed networks to expedite the generation of Real-World evidence (RWE) for regulatory decision making. In such networks, multiple data access providers (DAPs) collaborate by handling heterogeneous data and using the state-of-the-art approach to generate RWE. This involves the use of a common protocol, a common data model and common analytics with tools, which have been improving over the past 15 years.^3^ Comprehensive analyses of these approaches have been published.^4^

The lack of knowledge regarding the use and safety of medicines and vaccines during pregnancy and lactation is widely recognized. Studies indicate that 70-90% of women are exposed to a prescription medicine during pregnancy.^5^ However, the process of determining the teratogenic status of a novel medication currently takes 27 years.^6^ To close this evidence gap, the IMI ConcePTION project was established (www.imi-conception.eu), bringing together 88 stakeholders across Europe with the aim to create a learning healthcare ecosystem to support future studies on safety of medicines and vaccines during pregnancy and lactation. ConcePTION uses RWD to generate Real World Evidence (RWE) for childbearing-aged, pregnant, and lactating women, and their offspring.

Before conducting any analysis using RWD, it is imperative to ensure the quality of the data and Data Quality Assessments (DQAs) should be conducted to evaluate whether data sources are fit for purpose prior to study initiation. ConcePTION along with other research networks such as US Food and Drug Administration (FDA) Sentinel, PCORnet, and the Observational Health Data Science and Informatics program (OHDSI) perform DQAs on the data loaded into common data models (CDM) used by these networks for distributed analyses.^7–9^ The ConcePTION CDM allows for syntactic harmonization of heterogeneous data sources, and enables fitness-for-use and fitness-for-purpose evaluations.^10^ As part of the ConcePTION project, we have developed INSIGHT, a generic RWD tool to perform DQAs on ConcePTION CDM-standardized data sources ensuring data quality and the generation of reliable evidence for pharmacoepidemiologic studies. This tool aligns with Kahn’s data quality framework which encompasses three main quality dimensions: *conformance*, *completeness*, and *plausibility*.^11^

In this article, we describe INSIGHT, an open-source tool that comprises three levels of DQAs and we outline step-by-step its design and execution. Furthermore, we present an overview of the DQA workflow, including processes, visualizations, and responsible parties. We show examples of detected inconsistencies and errors, which serve as proof-of-concept, highlighting the tool’s effectiveness in identifying and addressing data quality issues.

## METHODS

As part of a common analytics and distributed analytics framework, Data Access Providers (DAPs) are required to convert their local data sources into the ConcePTION CDM using an Extract-Transform-Load (ETL) process. The ConcePTION CDM encompasses demographics, observation periods, medicines and vaccines exposure, events, procedures, mother-child linkage, visits, and clinical observations. The current version 2.2 includes 16 tables (Box 1). Additional details can be found elsewhere.^10^

#### Box 1.

**ConcePTION CDM tables**

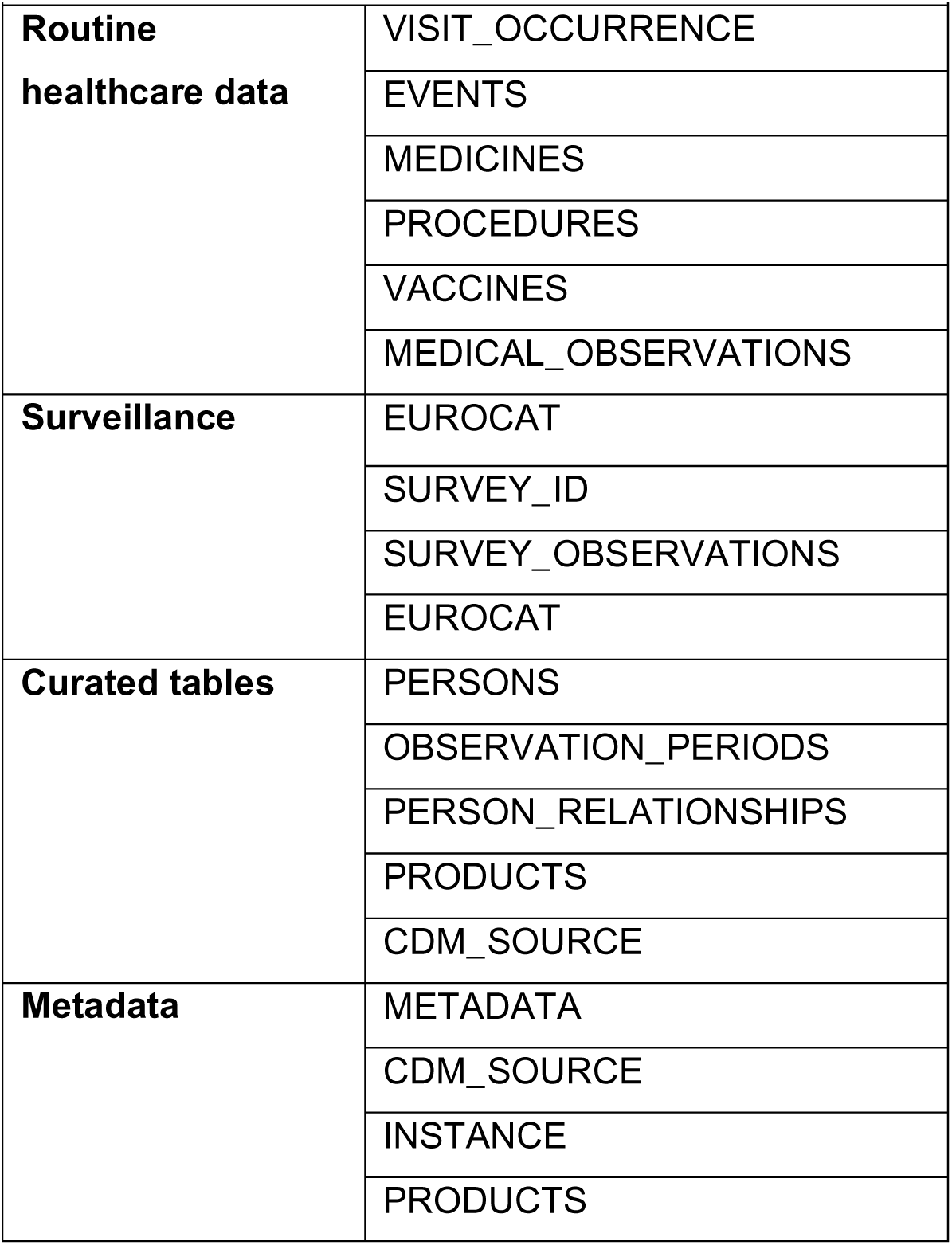

### ConcePTION Common data model

When working on studies with the ConcePTION CDM, DAPs are asked to load their data into a standardized data structure using the source values. This ensures that the ConcePTION CDM is syntactically (ensuring consistency in the structure) harmonized. The ConcePTION CDM pipeline does not require semantic harmonization (translation/mapping of source value to common vocabularies), which can often be time consuming. Instead, mapping is conducted as part of the study script, making it a transparent and flexible process. Once the data sources are syntactically standardized, DQAs are performed to allow assessment of data quality and fitness for purpose.

### Objectives of Data Quality Assessments

Various conceptual data quality frameworks have been published over the last years.^12^ For the development of INSIGHT, we focused on the harmonized terminology for data quality dimensions put forth by Khan et al., and the quality indicators established by the US FDA Sentinel initiative, OHDSI, and the EUROCAT indicators for population-based healthcare data sources.^11,13^ By incorporating these frameworks, we aimed to create a comprehensive and robust data quality verification pipeline for projects using the ConcePTION CDM. In Figure 1, we show the hierarchy of DQAs in INSIGHT followed by a brief description.

**Figure 1.**
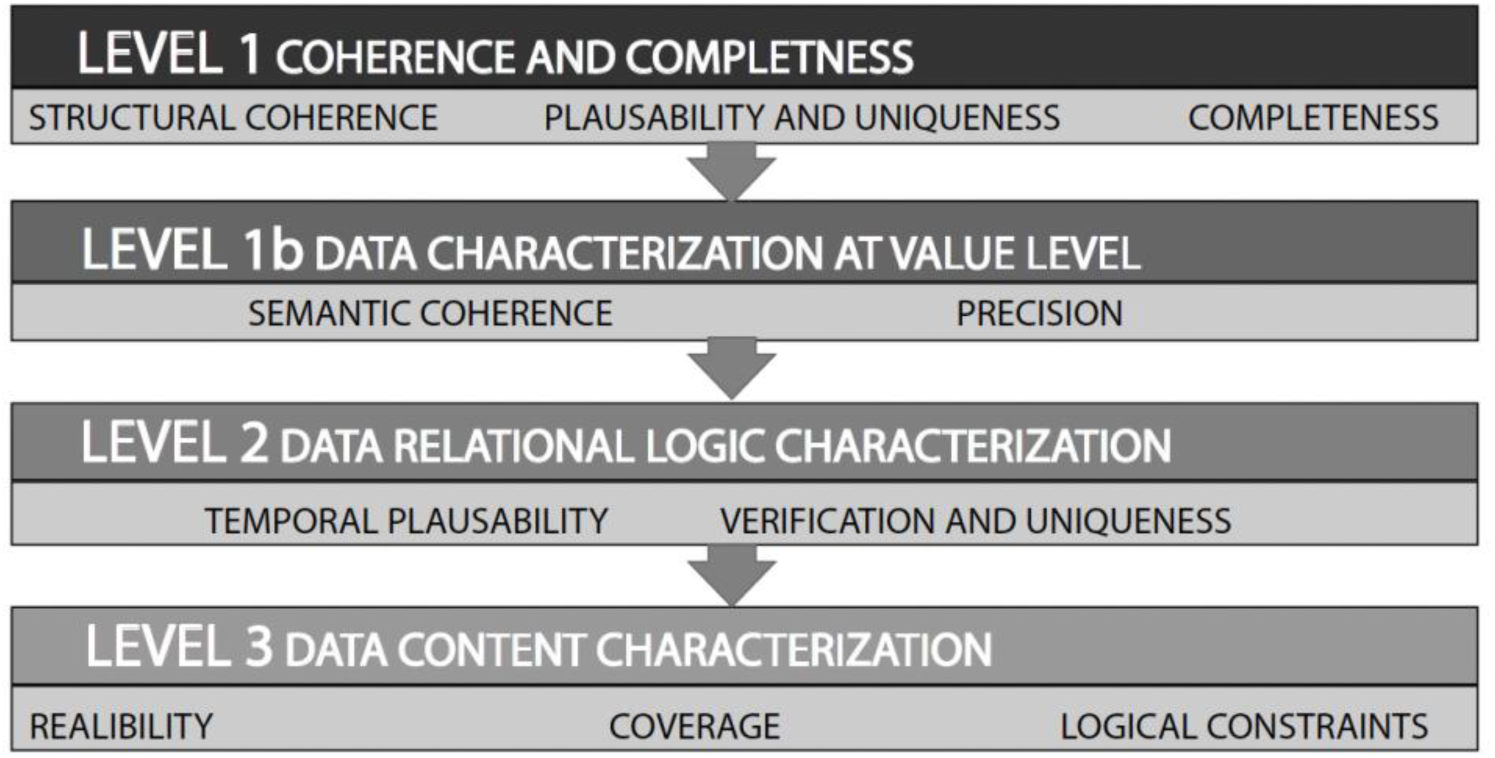
Hierarchy and dimensions of data quality assessment in INSIGHT

Level 1 data quality indicators provide insight on the completeness and compliance of the ETL process with the ConcePTION CDM specifications, ensuring format, structural and relational coherence. In addition, the plausibility of the data and the uniqueness of records is addressed, maintaining data integrity as well as identifying duplication errors.

Level 1b data quality indicators provide a list of all extracted and loaded non-date variables across the ConcePTION CDM tables. The primary objective is to provide DAPs, principal investigators, programmers, and analysts a comprehensive overview of the values of each non-date variable to allow for study-specific variable definitions. This level aims to enhance the understanding of the data content, facilitating semantic harmonization in the study scripts.

Level 2 data quality indicators provide an overview of the logical relationship and integrity of values within and between variables and tables. Its purpose is to verify the temporal plausibility by examining records that occur outside the recorded person-time, identifying observations related to a person ID that is not present in the PERSONS table, among other similar inconsistencies.

Level 3 data quality indicators provide distributions of population, diagnoses, medicines, vaccines, lifestyle factors, pregnancy, and temporal trends over calendar time for each specific variable. The primary objective is to allow for inspection of temporal changes in population, follow-up, medicines, vaccines, and disease rates. These indicators can be compared between instances and between DAPs, but also against external benchmarks to verify their fitness for purpose.

To ensure that data quality indicators can be inspected, it is important to present the results in a format that facilitates their understanding and sharing. To achieve this, HTML reports with the presentation of the data quality indicators were developed. These reports contain summary tables which allow for a concise representation of data quality indicators and graphs that provide a visual representation of trends and patterns. INSIGHT has been implemented using R scripts, which automate the running process of DQAs. The INSIGHT DQA is an iterative process, each level can be rerun until the required quality is attained or all constraints are noted.

### Design of Data Quality Indicators

#### Level 1: Coherence and Completeness

Level 1 comprises five major steps (Box 2). Step 1 verifies the presence and format of variables ensuring their inclusion and adherence to the designated format. Step 2 verifies missingness of values within the ConcePTION CDM and provides the extent of missing data overall and by calendar year. Step 3 verifies whether date variables comply with the required format and validates day, month, and year values. Steps 1 to 3 are performed simultaneously on all ConcePTION CDM tables, except for the METADATA table. The results are consolidated into a single HTML report.

#### Box 2.

**Details on the quality checks involved in level 1: step 0 to 5**

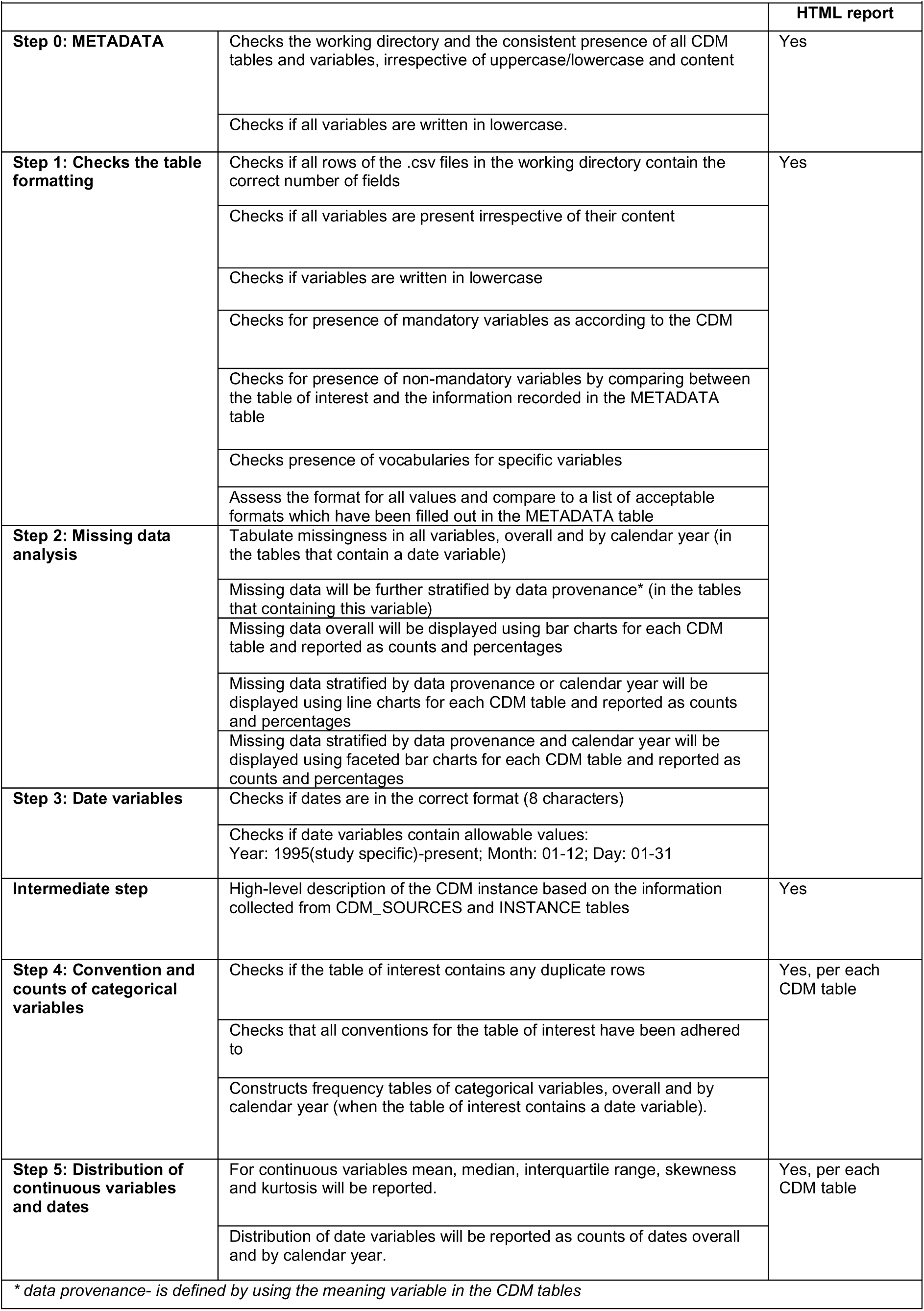

An intermediate step before Steps 4 to 5 generates a high-level characterization of the ConcePTION CDM instance by extracting information from the CDM_SOURCE and INSTANCE tables, highlighting the key attributes of the data instance. A verification of uniqueness of records is also performed as duplicates check. Step 4 verifies compliance of the ConcePTION CDM tables with the required conventions. These conventions are specific to each CDM table and refer to the table structure and vocabulary rules. Moreover, frequency tables are constructed for categorical variables, providing a summary of values. Step 5 produces the distribution of continuous variables and date variables. For continuous variables, descriptive statistics such as mean, median, interquartile ranges are calculated. Steps 4 and 5 are performed separately for each CDM table. An HTML report is generated for each table, providing a separate overview of the results. All issues identified need to be fixed by updating the ETL design, or when not possible will be noted. In summary, Level 1 DQA aims to verify format coherence and completeness of each variable across the 15 CDM tables, encompassing a total of 356 individual indicators. The code is available at https://github.com/UMC-Utrecht-RWE/INSIGHT-Level1

#### Level 1b: Data characterization at value level

Level 1b relies on the successful conformance to the ConcePTION CDM specifications and consists of two steps (Box 3). In this level, various aspects related to the data’s coherence and precision are evaluated through 26 checks (Box 3), including alignment of ATC codes at the same pharmacological level, or consistency of treatment duration expression (days or weeks) if available, among others. Level 1b does not generate an HTML report, but a set of csv files for each CDM table. The code is available at https://github.com/UMC-Utrecht-RWE/INSIGHT-Level1b

#### Box 3.

**Details on the quality checks involved in level 1b**

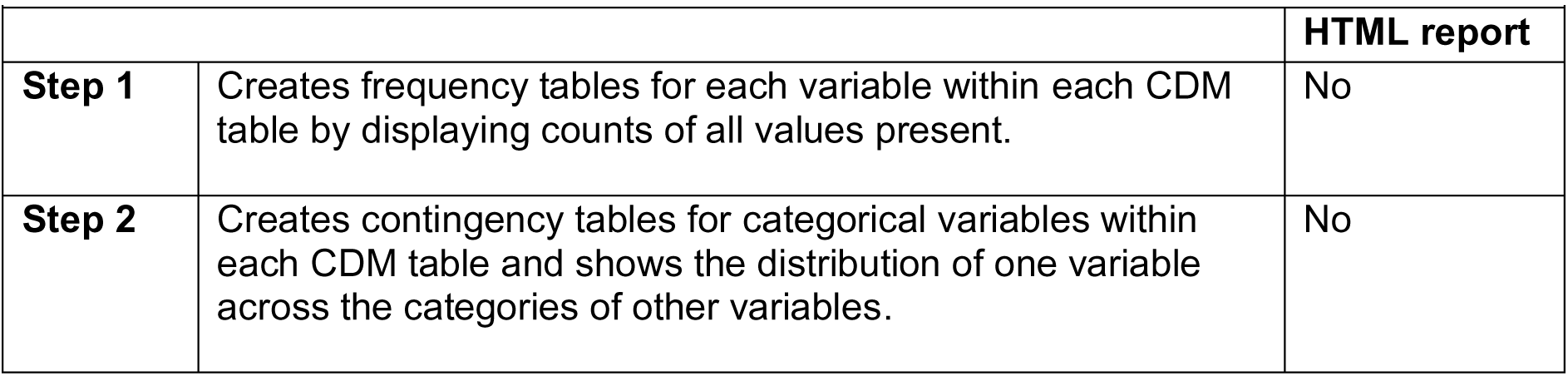

#### Level 2: Data Relational logic characterization

Level 2 consists of 8 steps (Box 4) with a total of 57 indicators and verifies temporal plausibility of date variables (Steps 2.1-2.3, 2.5-2.6) and consistency of encounter records across different tables (Steps 2.4, 2.7-2.8). For example, step 2.1 examines if health encounters occurred before an individual’s date of birth, ensuring the chronological integrity of the data. Similarly, Step 2.4 verifies that person IDs with a record in any of the CDM tables are also listed in the PERSONS table. The R-script is available at https://github.com/UMC-Utrecht-RWE/INSIGHT-Level2

#### Box 4.

**Details on the quality checks involved in level 2: step 1 to 8**

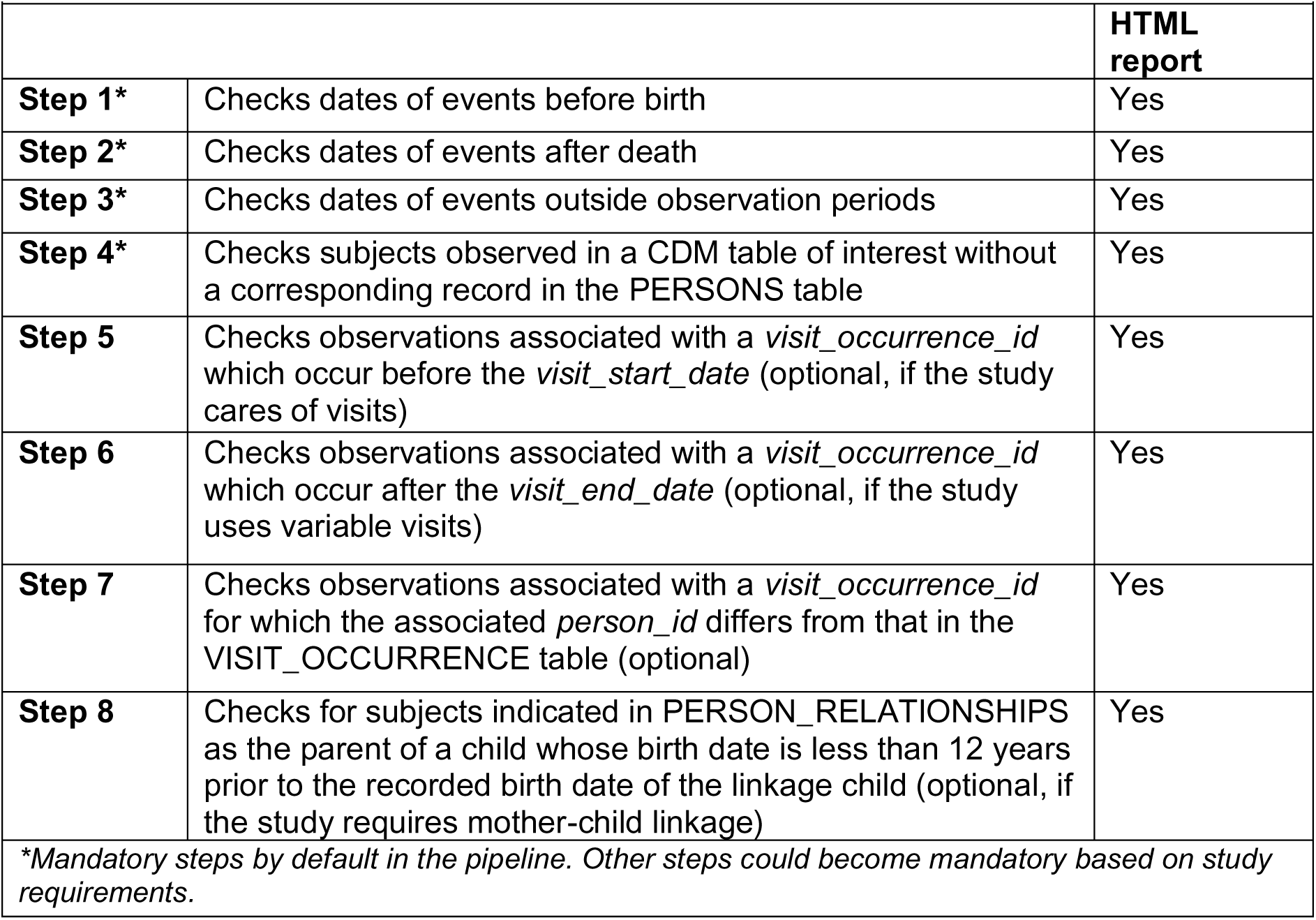

#### Level 3: Data Content Characterization

Level 3 creates tables and graphics for study-variables (Box 5) in 9 steps. First the population, its follow-up, and entry and exit patterns are described, allowing for verification of stability of the dynamic population. Subsequently key study variables such as medicines, vaccines, diagnoses, pregnancy, lifestyle factors and EUROCAT indicators are created in the study population. All indicators result in graphics allowing to inspect temporal trends, including person-time, prevalence of medicine prescriptions, and incidence of events for the study population and subpopulations of interest. This allows for the identification of any peaks, drops, or trends in the data and facilitates benchmarking between different instances, DAPs and external data. There are three versions of the output provided, one with no masking procedures applied, the two other versions allow for masking of cell counts. The first is masking all counts lower than five, and the second does not show absolute counts but only ranges. With over 249 indicators, Level 3 allows to deeply assess whether data is fit for purpose.The code is available at https://github.com/UMC-Utrecht-RWE/INSIGHT-Level3

#### Box 5.

**Details on the quality checks involved in level 3: step 1 to 9**

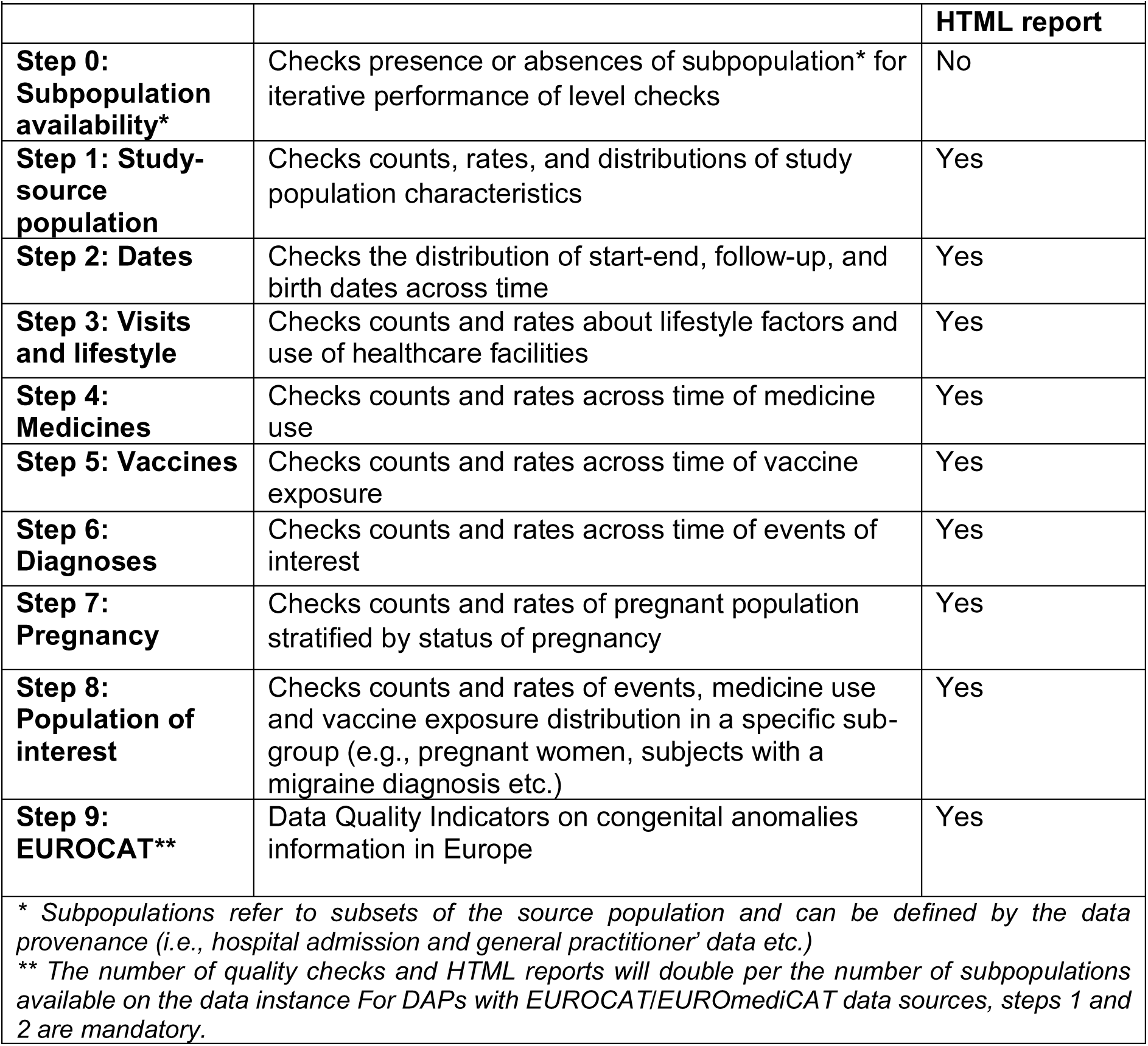

### HTML report

HTML reports are generated for Level 1, 2 and 3 outputs. Within Level 1 several reports are created. The first report lists the presence of the METADATA table, a mandatory table, in the working directory. The report on Step 1 to 3 provides information on the format coherence and completeness. Reports Step 4 to 5 are optional and based on study specific requirements of CDM tables (Box 1). Reports 1-4 in Level 2 are mandatory, whereas reports 5-8 are optional, depending on study requirements (Box 3). Level 3 DQAs produce nine reports of which report 1 and 2 are mandatory. The total number of reports depends on the study requirements and the number of subpopulations (Box 5). The output of level 1b is available as csv files and makes it possible to highlight any inconsistencies or create dictionaries for semantic harmonization of non-harmonized values.

### Responsible Parties

The R-scripts are available on the UMCU-RWE private GitHub and maintained by UMC Utrecht. A public frozen version is available for this paper. DAPs are responsible for downloading and running the R scripts locally and addressing issues related to table formatting that results from Level 1 Step 1 to 3 and exceeding the *a-priori* 5% inconsistencies threshold defined in Level 2. If any issues are reported, DAPs must revise the ETL of the data instance and rerun these steps.

The outputs of the Level 1, 1b, 2 and 3 DQAs are uploaded to the UMCU Digital Research Environment (DRE) by the DAPs, for centralized review by the data quality revisor and the DAPs.^14^ The data quality revisor collaborates with each DAP to address any issues flagged during the revision, as DAPs can provide background information on data collection processes, population characteristics, national healthcare system, availability of healthcare facilities, regulatory aspects, and expected disease incidence and prevalence. Observations are listed in the data quality approval form (Supplemental File 1). Data analytics engineers provide technical support related to issues that get reported. Figure 2 shows the workflow.

**Figure 2.**
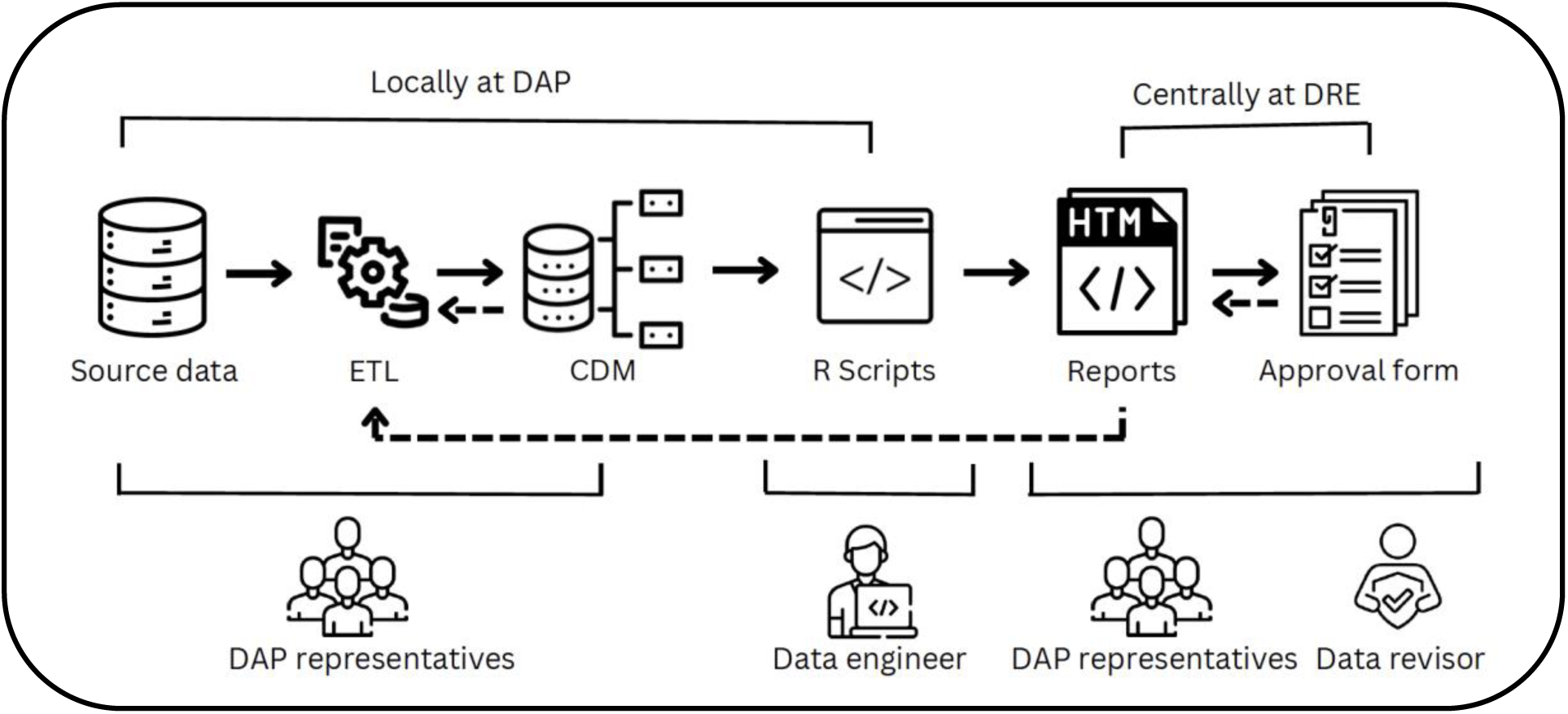
Workflow of INSIGHT

### Approval form

An approval form must be completed centrally by the DAP and the data quality revisor, with one form per instance and per level check. This form serves as a quality assurance document and systematically and transparently consolidates relevant information from the DQAs. The form is an Excel document with four tabs, one per level check listing the relevant aspects to be assessed per output (Supplemental File 1).

### Training

As VAC4EU utilizes the ConcePTION CDM pipeline for its vaccine evaluation studies, they have developed a training course dedicated to Data Quality checks. Investigators can register and complete this course online. Upon successfully passing the final test, they will be awarded certification.

## ILUSTRATIVE EXAMPLE

INSIGHT is available as a set of executable R scripts, currently supporting the use of ConcePTION CDM version 2.2.^15–18^

### Level 1

In Level 1, one of the key objectives is to assess conformance of the ETL’ed data to the ConcePTION CDM specifications. For instance, if alternative values such as “1” for males and “2” for females are used instead of the designated values (M, F, U, O) for the variable *sex_at_instance_creation* in the PERSONS table, it will be flagged as an error in Steps 1 to 3 report.

Additionally, Level 1 identifies unallowable values in date variables. In Figure 3, we show an example from the EVENTS table, the indicator *error_year* is displayed in blue, highlighting records with dates before 1995 or in the future. Most DAPs have data banks starting after 1995, that is why this cut off was chosen. The indicator *future_dates* show records with dates in the future in comparison to the script running date.

**Figure 3.**
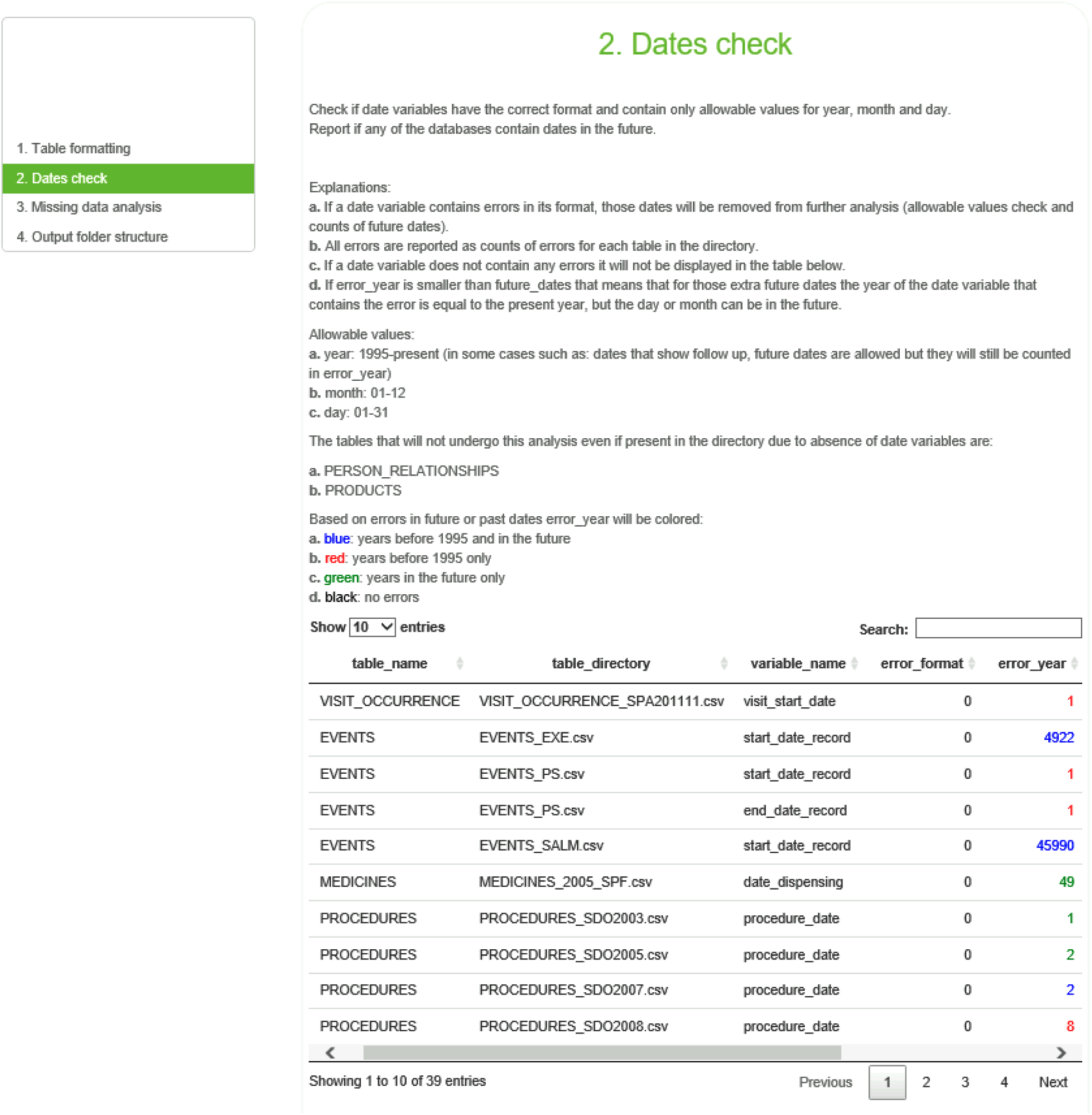
Example of level 1 check

DAPs are expected to resolve these issues by updating their ETL specifications to adhere to the specified rules and requirements, if possible.

### Level 1b

Level 1b allows users to visualize various aspects of the data, such as the composition of codes in the PROCEDURES table (e.g., number, number with letters, symbols like “$” or “?”) and provides insights into the original value definitions for study variables in the MEDICAL_OBSERVATIONS or SURVEY_OBSERVATIONS tables. Additionally, by assessing the content of the ConcePTION CDM tables the user can confirm that each table was fed appropriately.

### Level 2

Step 2.3 highlights records where dates fall outside the observation period (Figure 4). The construction of this period is a crucial step. Start dates of observation can be obtained from various sources, usually inhabitant registries. However, accessibility to these sources may vary and alternative resources such as first visit to a general practitioner are used. In figure 4, the high percentages of dates outside the observation period indicate that the current observation period fails to capture all relevant events.

**Figure 4.**
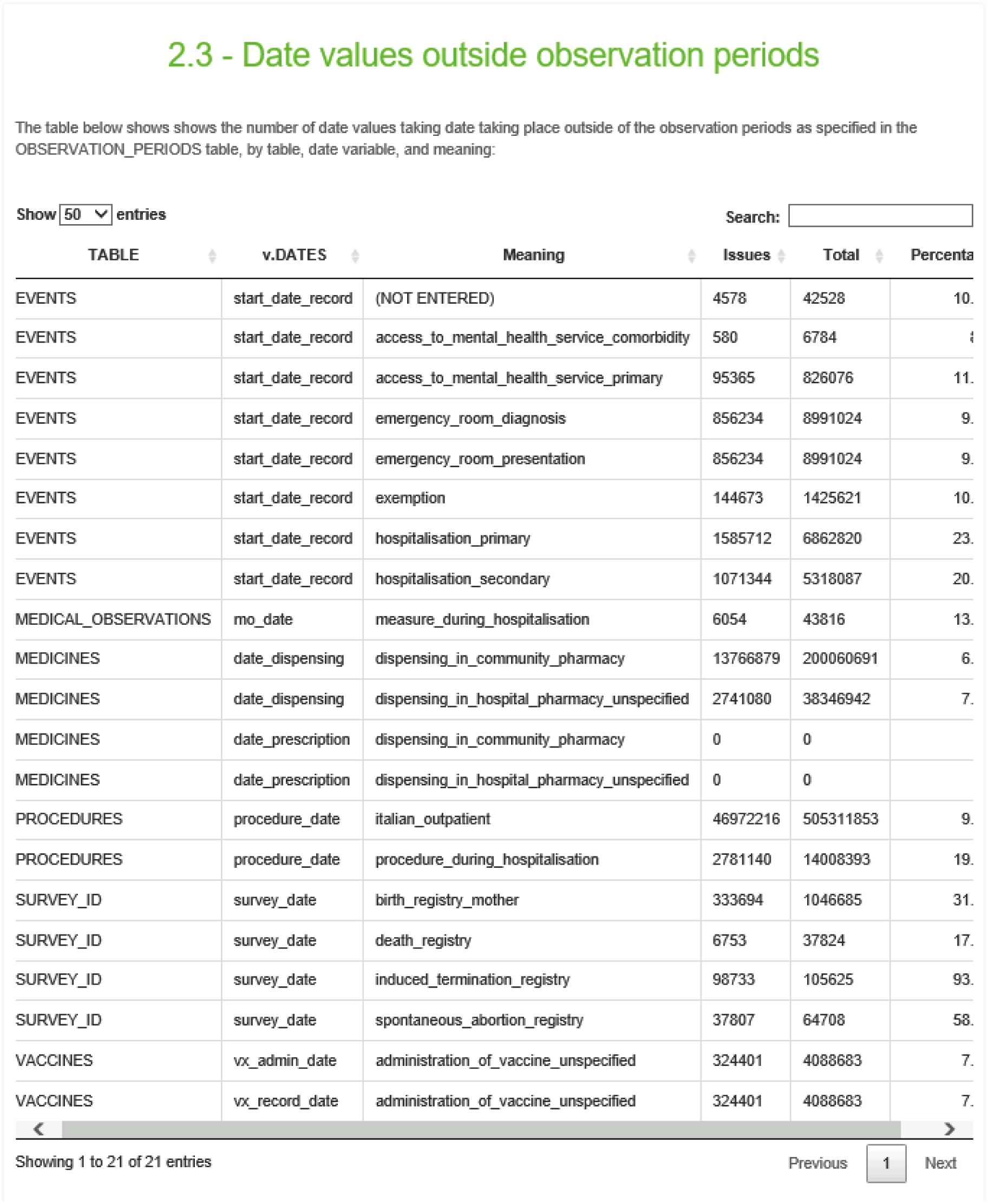
Example of level 2 check

Moreover, Level 2 assesses the presence of subjects across the CDM tables. Ideally, all *person_id* in the CDM tables should be in the PERSONS table. An error in step 4 indicates a need to re-evaluate the design of the PERSONS table.

### Level 3

In Level 3, DQAs become more extensive, as their goal is to facilitate the evaluation of fitness-for-purpose. Figure 5 illustrates a demographic tree indicating a similar distribution between females and males. However, the study focus of ConcePTION is on females and young males. It is possible that fathers have been included in the study population, which does not align with the study requirements but is not necessarily an error. Another example is gestational diabetes diagnostic codes in males. This could be explained by recording diagnoses under male children. However, this percentage should be low to reflect the reality of the condition, otherwise it may indicate an error.

**Figure 5.**
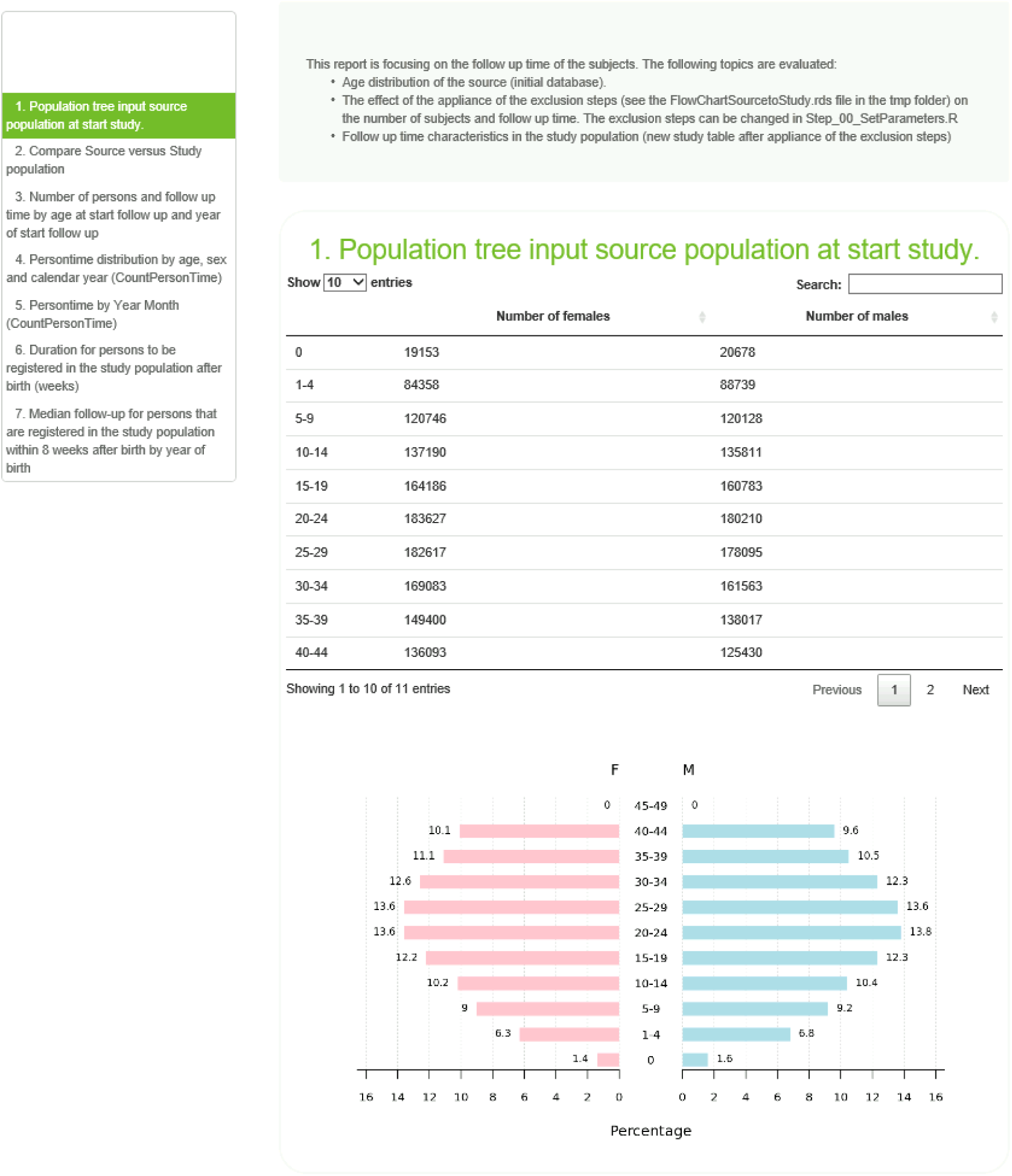
Example of level 3 check

In level 3, one of the checks is distribution analysis of start and end dates of follow-up, by year and month. In Figure 6 all records start in January and July of 2004. This pattern indicates a potential ETL specification to filter out records outside the study period of interest. The peak in July suggests imputation due to missing data in date of birth, which has been used as start of observation.

**Figure 6.**
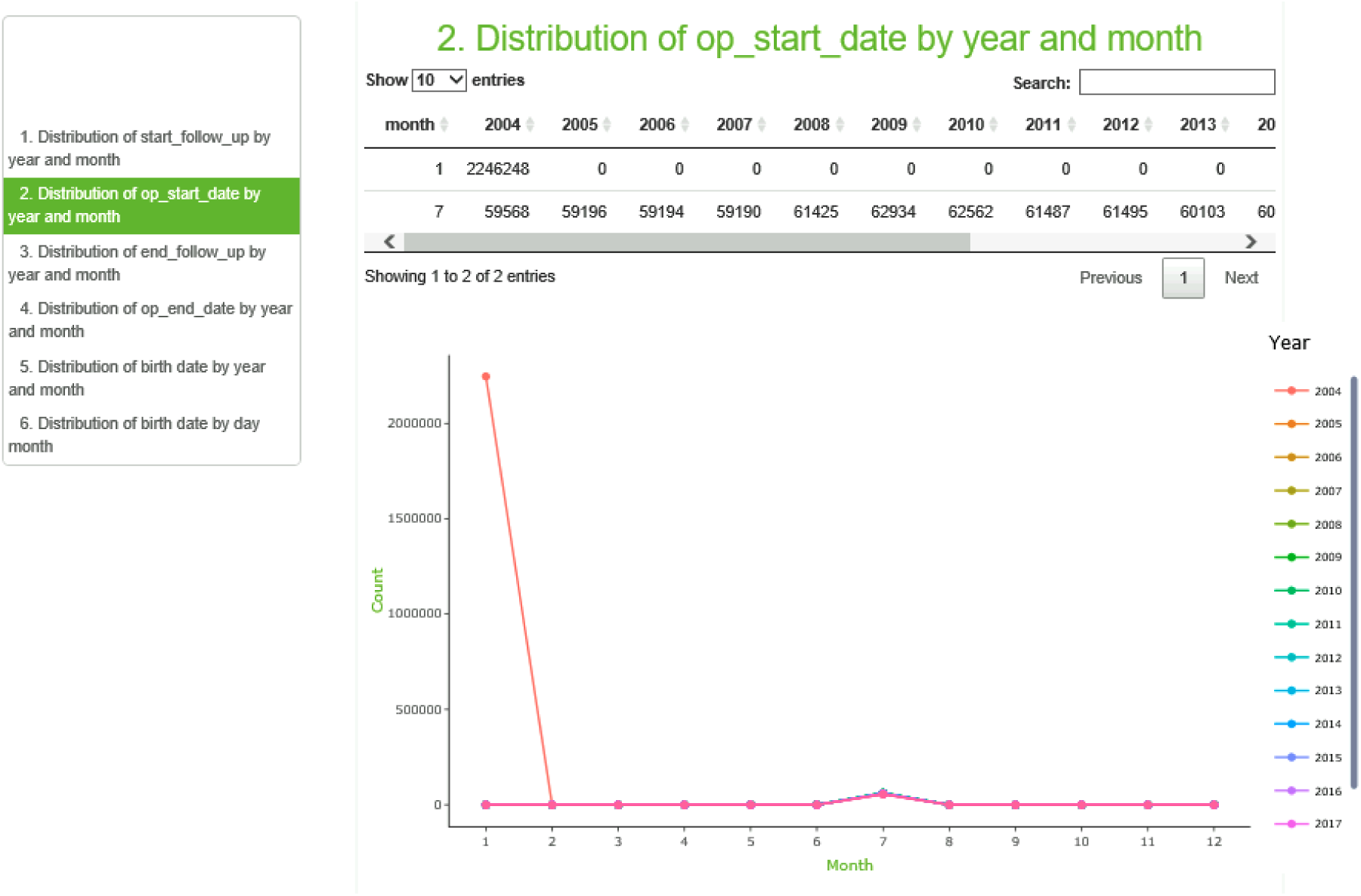
Example of level 3 check

Level 3 plays a crucial role in detecting and reflecting the consequences of technical errors during L1, L1b, and L2 and highlights any errors from the data generation processes. By identifying and addressing these issues, the goal is to improve the overall quality of the data source. INSIGHT is an iterative process of running and updating, until the DAPs and PI have confidence in the data source’s quality.

## DISCUSSION

The use of RWD in research and decision-making has gained significant attention from regulatory agencies such as FDA, PMDA, and EMA. They have emphasized the importance of valid and trustworthy RWD for generation of RWE.^1,2^ In this article, we present INSIGHT, a tool to assess fit-for-use (research independent) and fit-for-purpose (research specific) of RWD.

Studies including multiple data sources are becoming more popular especially in pharmacoepidemiologic studies.^19^ With the abundance of healthcare data available, DAPs can leverage various data sources including routine healthcare data, surveillance systems, inhabitant registries, and more. However, the utility of these granular data sources relies on their quality. Data quality encompasses several features, including representativeness, precision, and accessibility, among other dimensions. To evaluate quality, EMA and Heads of Medicines Agency have proposed the DQF for EU medicines regulation which includes five quality dimensions: reliability, extensiveness, coherence, timeliness, and relevance.^20^ We have mapped INSIGHT’s data quality dimensions to Kahn’s framework (left), and the EMA DQF (right) (Supplemental File 2). INSIGHT is built on the ConcePTION CDM, which is utilized by multiple networks (ConcePTION project, VAC4EU, EU & PV network, SIGMA consortium) and in many regulatory required multi-data source studies commissioned or requested by EMA.

CDMs play a vital role in facilitating data quality assessment when using secondary-use data.^21^ While DQAs aid assessing fitness-for-purpose, it is essential to recognize that data quality is influenced by numerous factors, mostly the health care system, the purpose of data recording and the type of data recorder. INSIGHT DQAs provide a generic and transparent overview of data quality indicators, aiming to identify inconsistencies in the data, which can be categorized into three main types: extraction or convention errors, script errors, and patterns for acknowledgment and documentation. The outputs support the study team to understand the data that is being utilized and the semantic harmonization. The purpose of DQAs varies across these contexts, ranging from rectifying extraction errors to fine-tuning scripts to suit project requirements. Additionally, documenting data patterns ensures accurate interpretation and understanding of data nuances and limitations.

### Previous data quality assessment tools

Various research networks have developed their own DQA processes with varying levels and indicators. For example, the US FDA Sentinel uses a four-level approach focusing on completeness and validity, accuracy and integrity, consistency of trends, and plausibility.^9,22^ The EUROCAT network, has a comprehensive DQA process, that includes indicators such as case ascertainment, accuracy of diagnoses, data completeness, timeliness, and availability of denominator data.^23,24^ Researchers using the OMOP CDM have access to tools like the White Rabbit and the Data Quality Dashboard (DQD) for assessing data quality at source data and CDM-standardized database. ^8,25–27^

A comparison of DQAs across six research networks revealed variations in the number of quality checks, ranging from 875 up to 3234.^28^ The DQAs processes also varied, including differences in centralization, distributed coordination of DQAs, programing languages, and staff involvement. These differences can be attributed to specific network requirements, analytical focus, as well as the maturity of their DQA models.

### Strengths and limitations

Our open-source DQA methodology was built during the ConcePTION project and has been made generic and is adopted by other networks using the ConcePTION CDM. Our approval system is based on a flexible approval form and relies on study requirements, over predetermined conditions. However, there are limitations to be acknowledged. First, script running time depends on hardware, data size and complexity. Components like the central processing unit, random access memory, and computer data storage help determine process efficiency. Second, the pipeline requires iterative reruns after data issues are fixed until desired quality is achieved or at least, explained. Steps must be executed sequentially to avoid downstream issues. Last, fit-for-purpose evaluation is time and resource-intensive. While summary reports and dashboards aid assessment, automated detection of trends and verification against external benchmarks need to be facilitated further. Automating outlier and pattern detection is an area for future development given the increase complexity and limited skilled human resources. Overall, the DQA workflow is subject to continuous improvements.

### Implications for researchers and future research

The INSIGHT R scripts are openly available through GitHub, along with a training course for the Vaccine Monitoring Collaboration for Europe (VAC4EU) and educational videos to encourage other stakeholders to adopt this DQA pipeline and adhere to principles of open science. This tool has been already successfully implemented in EMA tendered risk minimization studies (EUPAS31095, EUPAS21001), COVID vaccines effectiveness study (EUPAS40404, EUPAS42504), Post-Authorization Safety Studies (EUPAS43593, EUPAS44424, EUPAS41725, EUPAS45461), and CONSIGN (EUPAS39226, EUPAS39438, EUPAS40317).^29^ As previously stated, INSIGHT is compliant with the proposed DQF for EU medicines regulation launched in October 2022.

Interoperability between CDMs and their specific DQAs processes remains a challenge, as each CDM must be used in conjunction with its customized tools. This implies that applying external DQAs tools to an already CDM-standardized data source would require an additional ETL process tailored to that CDM. To address this, we propose to establish standards and guidelines for achieving interoperability across different CDMs. This would facilitate the exchange of DQAs tools and methodologies between research networks and regulatory bodies, streamlining the quality assessment process.

## CONCLUSION

In this article, we introduced INSIGHT, a comprehensive pipeline for conducting DQAs of ConcePTION CDM-standardized datasets. The tool aims to facilitate the evaluation of the fitness-for-use and fitness-for-purpose of RWD. By implementing this tool, researchers can assess the adequacy and reliability of the data, ensuring that the evidence generated is robust and reliable for studies on medicines and vaccines. INSIGHT is a valuable tool for evaluating the quality of data sources and enhancing the credibility of RWD in clinical and regulatory decision-making.

## Data Availability

All data produced in the present study are available upon reasonable request to the authors

## AUTHORS CONTRIBUTION

VH, CD, and MCJM started with the initial design; VH, RE, EA, and RvdB programmed the R scripts; VH and CLAN wrote the initial draft of the manuscript. VH, CLAN, JRA, RE, EA and MCJM, provided critical feedback and edited the manuscript. All authors have seen and approved the manuscript. The research leading to these results was conducted as part of the ConcePTION consortium. This paper only reflects the personal views of the stated authors.

## ACKNOWLEDGMENTS

We would like to thank Rutger van den Bor for supporting the programming of INSIGHT Level 2, Claudia Bartolini for testing the scripts, and Rosa Gini for her expertise and collaboration throughout this project.

## SUPPLEMENTARY MATERIAL

**S1.** Approval form

**S2.** Theoretical mapping of INSIGHT to other DQFs

## INSIGHT PIPELINE AVAILABILITY

The quality check pipeline for ConcePTION CDM v2.2 instances is available as an open-source R scripts in the following link https://github.com/UMC-Utrecht-RWE. Protocol and Statistical Analysis Plan (SAP) can also be found there. Furthermore, a training course for DAPs and researchers is available upon request.

## STUDY REGISTRATION

This research was registered in EU PAS registration with number EU50142.

