## Supplementary figures and images for "INSIGHT: A Tool for Fit-for-Purpose Evaluation and Quality Assessment of Observational Data Sources for Real World Evidence on Medicine and Vaccine Safety"

### Supplemental File 2

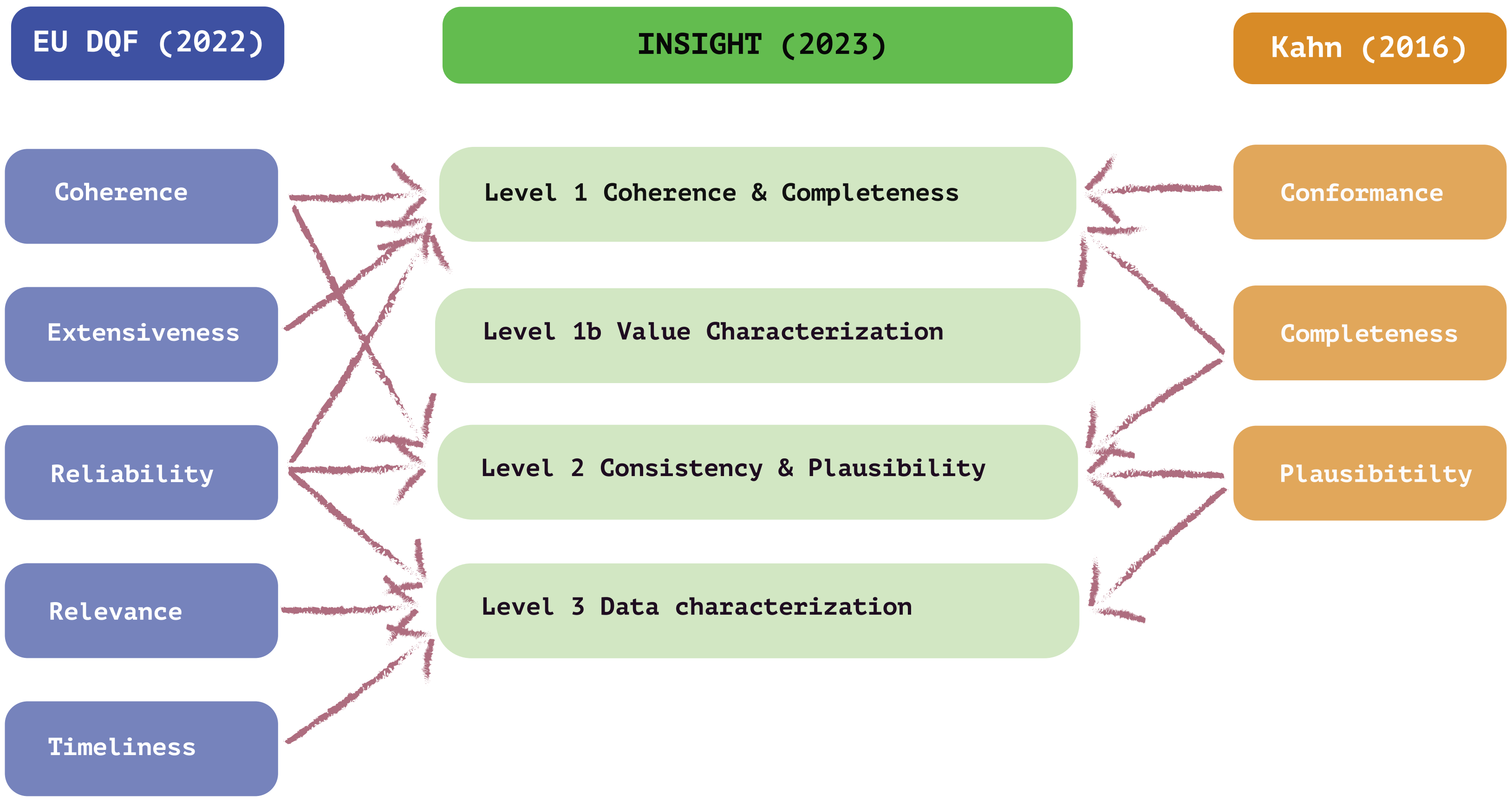
